# Trends in lipid-modifying agent use in 83 countries

**DOI:** 10.1101/2021.01.10.21249523

**Authors:** Joseph E Blais, Yue Wei, Kevin KW Yap, Hassan Alwafi, Tian-Tian Ma, Ruth Brauer, Wallis CY Lau, Kenneth KC Man, Chung Wah Siu, Kathryn C Tan, Ian CK Wong, Li Wei, Esther W Chan

## Abstract

**Aims:** Lipid-modifying agents (LMAs) are increasingly used to reduce lipid levels and prevent cardiovascular events, but the magnitude of their consumption in different world regions is unknown. We aimed to describe recent global trends in LMA consumption and to explore the relationship between country-level LMA consumption and cholesterol concentrations.

**Methods and results:** This cross-sectional and ecological study used monthly pharmaceutical sales data from January 2008 to December 2018, for 83 countries from the IQVIA Multinational Integrated Data Analysis System, and total and non–high-density lipoprotein (non– HDL) cholesterol concentrations from the NCD Risk Factor Collaboration. The compound annual growth rate (CAGR) was used to assess changes in LMA consumption over time. From 2008 to 2018, the use of LMAs increased from 7,468 to 11,197 standard units per 1000 inhabitants per year (CAGR 4.13%). Statins were the most used class of LMA and their market share increased in 75% of countries between 2008 and 2018. Fibrates were the most consumed nonstatin drugs. From 2013 to 2018, consumption of low-density lipoprotein lowering therapies increased (statins 3.99%; ezetimibe 4.01%; proprotein convertase subtilisin/kexin type 9 (PCSK9) inhibitors 104.47%). Limited evidence supports a clear relationship between country-level changes in LMA consumption and total and non–HDL cholesterol concentrations in 2008 versus 2018.

**Conclusion:** Since 2008, global access to LMAs, especially statins, has improved. In line with international lipid guideline recommendations, recent trends indicate growth in the use of statins, ezetimibe, and PCSK9 inhibitors. Country-level patterns of LMA use and total and non–HDL cholesterol varied considerably.

## INTRODUCTION

Lipid-modifying agents are widely recommended to reduce atherogenic lipid levels and to prevent atherosclerotic cardiovascular disease.(1–3) Over the last few decades, emerging evidence of the benefits and risks of different classes of lipid-modifying agents has informed updates to cholesterol guidelines and influenced treatment choices. Recommendations for the broader use of statins in the 2013 American College of Cardiology/American Heart Association (ACC/AHA) cholesterol guideline(4) raised concerns about extrapolating the available evidence, and subsequent worldwide ‘statinization’.(5) It is unknown whether overall lipid-modifying agent use has continued to increase at the global level. Given the robust evidence of benefit for statins, their share of lipid-modifying agent use may have evolved over time.

Several developing regions now face a greater burden from elevated cholesterol levels, yet they have lower consumption of lipid-modifying agents as compared to high-income regions.(6,7) Since 1980, parts of Africa and East and Southeast Asia have experienced increases in total and non–HDL cholesterol, while high-income Western countries have experienced decreases.(6) It is unknown whether recent consumption trends of lipid-modifying agents correspond with the observed changes in cholesterol levels. To date, there is also limited data on the use of lipid-modifying agents for many countries outside of North America and Europe.

In this study, we analyzed pharmaceutical market intelligence data from the IQVIA Multinational Integrated Data Analysis System (MIDAS) from 2008 to 2018. We aimed to describe contemporary trends in lipid-modifying agent use and to explore the relationship between the use of lipid-modifying agents and average total and non–HDL cholesterol concentrations. Regional and cross-country comparisons on trends of lipid-modifying agent consumption can be used to monitor progress on access to lipid-modifying treatment, to explore reasons for different patterns of use, and to connect information about an important cause of cardiovascular disease and its pharmacological treatment.

## METHODS

### Study Design and Data Sources

We conducted a cross-sectional and ecological study using monthly pharmaceutical sales data collected by IQVIA in 83 countries from January 2008 to December 2018. Country-level data is collected from multiple distribution channels, such as wholesalers, direct distribution from manufacturers, retail, and hospital pharmacies and differs by country and data type. If required, the sampled data is then projected to represent 100% of the total sales channel in each country. Product information in MIDAS is classified according to Anatomic Therapeutic Chemical (ATC) codes, and undergoes various quality-control checks. The available data for this study provides an international view of medication use across 39 high-, 24 upper-middle, 13 lower-middle, and 7 low-income economies included in 67 MIDAS country panels (Supplementary Table 1). MIDAS countries can be defined as individual countries, regions or territories (e.g., Puerto Rico), or groups of countries (i.e., Central America and French West Africa). IQVIA annually validates its data using manufacturer sales to estimate the accuracy of its data, with 95% global precision in most years.(8,9) Data from MIDAS has been used to assess global patterns in medicine consumption.(7,10) No information about patients or prescribers was available in our data; therefore, this study was exempt from ethical approval by the Institutional Review Board of the University of Hong Kong/Hospital Authority Hong Kong West Cluster.

We also used the mid-year population estimates from the United Nations (UN) World Population Prospects 2019 report.(11) The population of the constituent countries for French West Africa and Central America were summed for each year. Countries were then classified according to the UN geographical region (i.e. Africa, Latin America and the Caribbean, Oceania, Europe, Northern America, and Asia) and subregions. Country-level estimates for mean total and non–HDL cholesterol were obtained from the NCD Risk Factor Collaboration.(6) The NCD Risk Factor Collaboration used publicly available data from population-based surveys, and estimated age-standardized total and non-HDL cholesterol for 200 countries, from 1980 to 2018, using a Bayesian model with the Markov chain Monte Carlo algorithm.

### Lipid-modifying Agent Categories

The available data included sales of brand, generic, and combination lipid-modifying agents. Because of limited coverage by the WHO, IQVIA does not assign ATC codes or defined daily doses to combination products (e.g. *C10B Lipid modifying agents, combinations*).(12) In addition, because the WHO ATC system groups ezetimibe, PCSK9 inhibitors, and omega-3 fatty acids, under the *C10AX Other lipid modifying agents* category, we classified lipid-modifying agents into the following therapeutic categories: statins, fibrates, bile acid sequestrants, niacin, ezetimibe, omega-3 fatty acids, PCSK9 inhibitors, and others (Supplementary Table 2).

### Consumption Measurements

The primary variables of interest were the monthly number of standard units, and consumption standardized for population, defined as standard units per 1000 inhabitants, either per quarter or per year. The measure of standard units captures the entire market of products in MIDAS and includes combination products. IQVIA defines a standard unit as the smallest common dose which allows for comparisons between products of different dosage forms. For example, one standard unit equals a tablet, a capsule, or a pre-filled syringe (for PCSK9 inhibitors, for example).

### Data Analysis

We reported the total number of standard units sold per month for all lipid-modifying agents and by each drug class. For each calendar year from 2008 to 2018, monthly sales data were aggregated per quarter (Q1 = Jan, Feb, March; Q2 = April, May, June; Q3 = July, August, September; Q4 = October, November, December) and per year (January to December), and standardized per 1000 inhabitants. Consumption of all lipid-modifying agents by region and subregion was plotted. For all lipid-modifying agents and each class, we assessed consumption trends using the compound annual growth rate (CAGR). CAGR is the geometric mean of annual growth rates and is used to compare growth trends over several years and is defined as 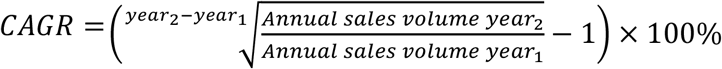.(13) Because CAGR is sensitive to the start and end years of the time period, we calculated it both for the overall study period (2018 vs 2008; or the earliest year of available data), and for the last five years of the study (2018 vs 2013). The latter five-year period is when most MIDAS country panel additions were completed and can capture consumption trends since the publication of the 2013 American College of Cardiology/American Heart Association cholesterol guideline (Supplementary Table 1).(4) We estimated CAGR for PCSK9 inhibitors by comparing the annual sales in 2016 and 2018, since evolocumab and alirocumab were first approved in July 2015, and thus did not have a 12 months of sales data in 2015.(14,15)

Statin market share was estimated as the percentage of statin standard units sold relative to all lipid-modifying agent standard units in 2008 versus 2018. We plotted lipid-modifying agent consumption versus sex-specific cholesterol concentrations in 2018 and assessed their correlation using Spearman’s *pp*. Furthermore, we plotted the change in annual lipid-modifying agent consumption and mean concentrations of total cholesterol and non–HDL cholesterol for 2008 to 2018. For these plots, we excluded countries with no consumption data in 2008 (i.e. Thailand, Bosnia, Kazakhstan, Serbia, and Netherlands). Furthermore, for the MIDAS countries of French West Africa and Central America, we selected the mean cholesterol concentrations for Togo and El Salvador, respectively, to represent changes for these MIDAS countries. These countries were arbitrarily selected since their cholesterol concentrations were approximately in the middle of the plots for average cholesterol in their respective MIDAS country. Two authors (JEB and YW) independently accessed the data and conducted the analysis using R software version 3.6.1 (R Core Team; Vienna, Austria).

## RESULTS

### Consumption Volume

Use of all lipid-modifying agents increased from a total of 2,904,258,689 standard units in January 2008 to 5,509,412,107 in December 2018. Statins were the most commonly used class of lipid-modifying agent (Figure 1A). Fibrates, ezetimibe, and omega-3 fatty acids were the most consumed nonstatins drugs (Figure 1B). When accounting for the population of the included countries, the annual use of lipid-modifying agents increased at a CAGR of 4.13% from 2008 to 2018. In the latter five-year period (2013-2018), consumption slowed with a CAGR of 3.16% (Table 1). Statins experienced the largest average growth from 2008 to 2018 (CAGR 5.19%). Between 2013 to 2018, trends for nonstatins differed. There was increased consumption of other lipid-modifying agents, ezetimibe, and PCSK9 inhibitors. In contrast, consumption of fibrates, niacin, omega-3 fatty acids, and bile acid sequestrants decreased (Table 1).

**Table 1.** Use and Growth Rate of Lipid-Modifying Agents by Category in 83 Countries Between 2008 to 2018

|  | Consumption (standard units per 1000 inhabitants) |  |  |  |  |  |  |  |  |  |  | CAGR (%) |  |
| --- | --- | --- | --- | --- | --- | --- | --- | --- | --- | --- | --- | --- | --- |
|  | 2008 | 2009 | 2010 | 2011 | 2012 | 2013 | 2014 | 2015 | 2016 | 2017 | 2018 | 2008-2018 | 2013-2018 |
| All lipid-modifying agents | 7,467.86 | 8,089.53 | 8,694.76 | 9,149.65 | 9,289.57 | 9,585.08 | 9,839.76 | 10,064.89 | 10,464.85 | 10,767.08 | 11,197.07 | 4.13 | 3.16 |
| Statins | 5,706.76 | 6,261.74 | 6,761.64 | 7,223.41 | 7,434.18 | 7,780.26 | 8,088.59 | 8,367.84 | 8,777.62 | 9,071.05 | 9,463.05 | 5.19 | 3.99 |
| Fibrates | 710.33 | 755.87 | 825.22 | 824.99 | 815.49 | 821.09 | 823.63 | 812.87 | 800.15 | 781.88 | 774.58 | 0.87 | -1.16 |
| Ezetimibe | 482.86 | 439.23 | 422.83 | 411.14 | 402.27 | 400.70 | 390.03 | 386.46 | 410.38 | 441.26 | 487.67 | 0.10 | 4.01 |
| Omega-3 fatty acids | 237.89 | 282.92 | 311.55 | 320.67 | 309.58 | 276.16 | 256.37 | 245.67 | 236.51 | 237.44 | 241.83 | 0.16 | -2.62 |
| Niacin | 210.98 | 221.82 | 243.05 | 241.01 | 205.69 | 180.94 | 159.89 | 135.73 | 128.18 | 128.16 | 126.45 | -4.99 | -6.92 |
| Bile acid sequestrants | 103.72 | 112.50 | 115.15 | 112.17 | 105.27 | 107.34 | 100.13 | 93.84 | 87.03 | 80.56 | 79.69 | -2.60 | -5.78 |
| PCSK9 inhibitors | 0.00 | 0.00 | 0.00 | 0.00 | 0.00 | 0.00 | 0.00 | 0.02 | 0.13 | 0.32 | 0.56 | NA | 104.47<sup>a</sup> |
| Others | 15.32 | 15.45 | 15.33 | 16.26 | 17.09 | 18.58 | 21.12 | 22.47 | 24.85 | 26.41 | 23.23 | 4.25 | 4.57 |

<sup>a</sup>CAGR calculated from 2016-2018.
Abbreviations: CAGR, compound annual growth rate; NA, not applicable.

**Figure 1.**
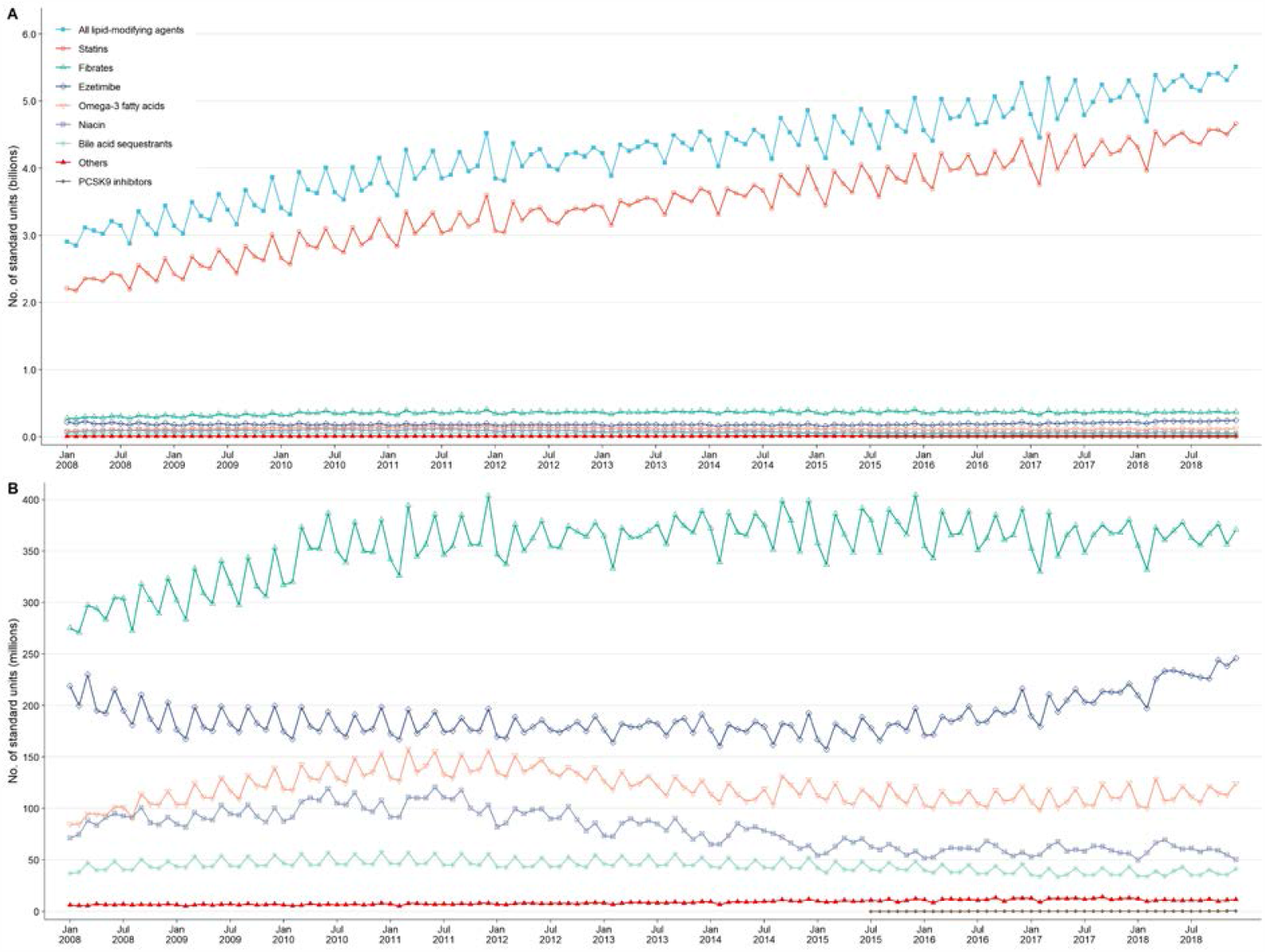
Monthly Number of Lipid-Modifying Agent Standard Units Used in 83 Countries from 2008 to 2018. Monthly sales data are from IQVIA MIDAS. Lipid-modifying agents are categorized according to therapeutic categories (see Supplementary Table 2). **A**, number of standard units (billions) sold of all lipid-modifying agents and according to therapeutic category. **B**, number of standard units (millions) sold of fibrates, ezetimibe, omega-3 fatty acids, niacin, bile acid sequestrants, other lipid-modifying agents, and PCSK9 inhibitors.

The use of all lipid-modifying agents over time varied widely according to region, subregion, and country. The overall use of lipid-modifying agents in Northern America remained similar in 2018 as compared with 2008, while other regions saw steady increases in consumption (Figure 2A). In 2018, lipid-modifying agent consumption in Oceania was nearly 7.3 times greater than Asia, 6.1 times greater than Latin America and the Caribbean, and 16.1 times greater than Africa. At the subregion level, Central Asia, Central America, and Western Africa had the lowest use of lipid-modifying agents (Figure 2B). In 2008, use of lipid-modifying agents in countries outside Northern America and Europe was low (Figure 3A). By 2018, use had increased in most countries (Figure 3B), notably in South America, Southern Europe, Northern Africa, and in East Asia.

**Figure 2.**
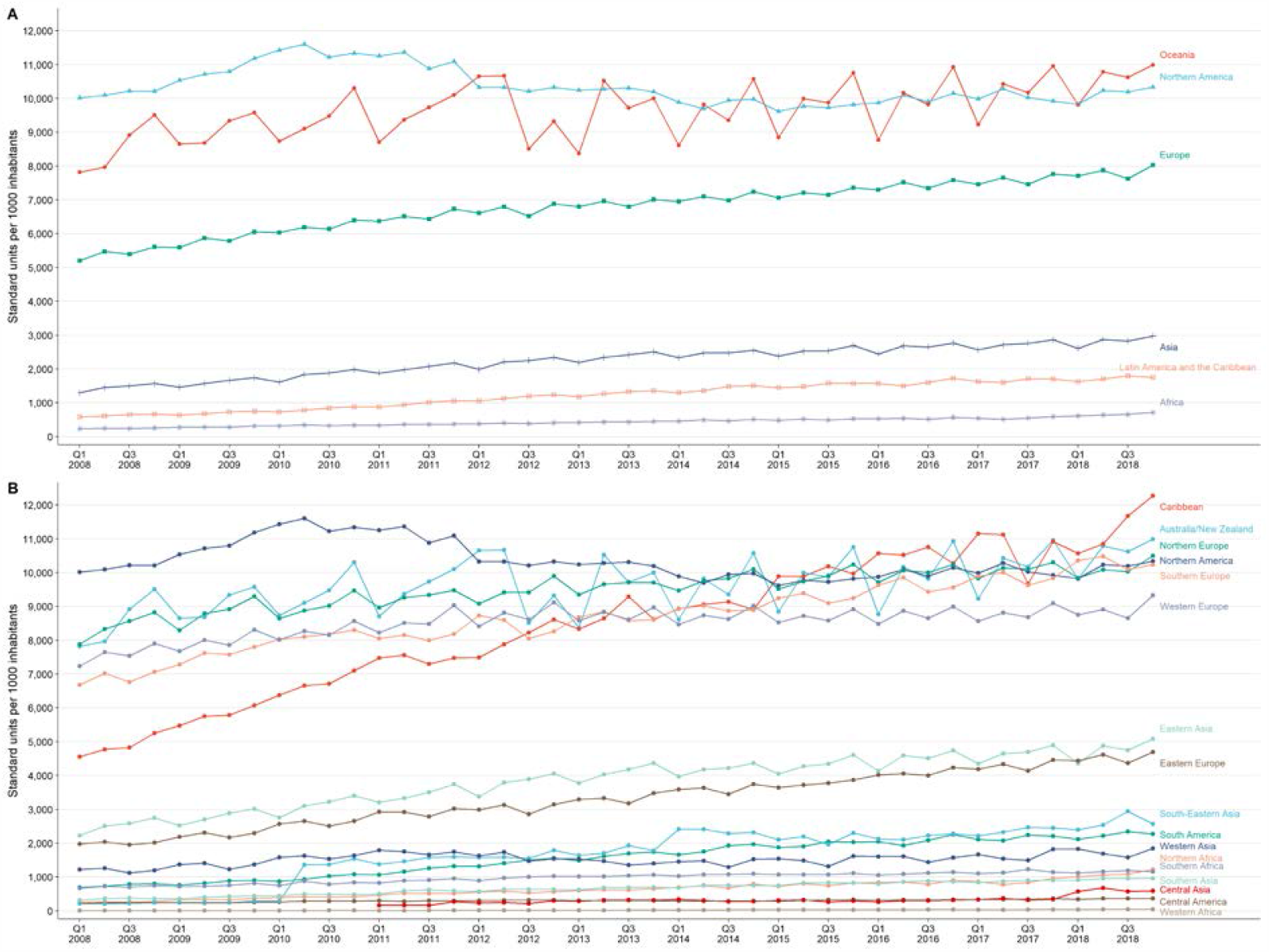
Quarterly Use of Lipid-Modifying Agents by Region from 2008 to 2018. Monthly sales data were aggregated on a quarterly basis. Individual countries were grouped according to UN geographical regions and subregions (see Supplementary Table 1).

**Figure 3.**
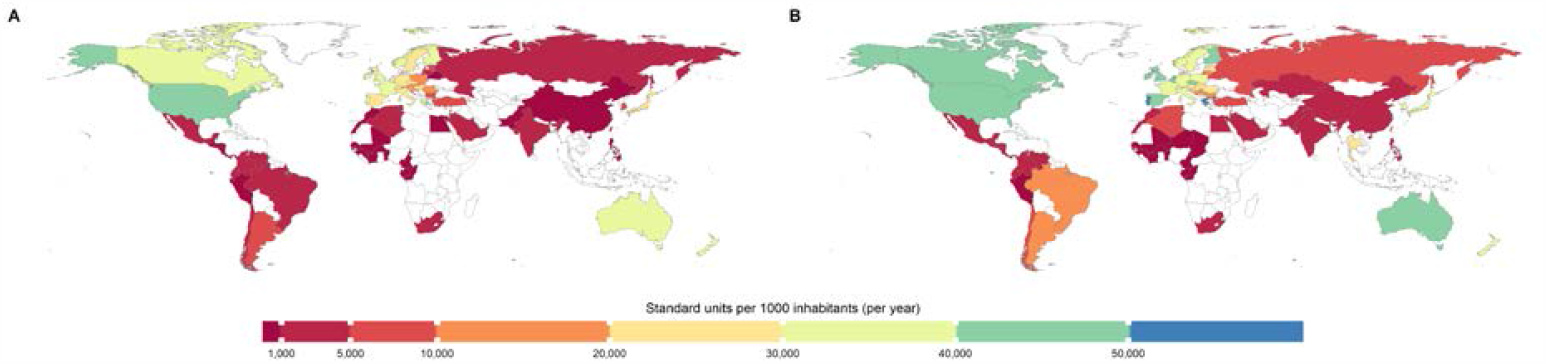
Use of Lipid-Modifying Agents by Country in 2008 and 2018. **A**, consumption of lipid-modifying agents in 2008. **B**, consumption of lipid-modifying agents in 2018.

Countries in Northern America and Europe were the greatest consumers of lipid-modifying agents. In 2008, the countries with the highest consumption of lipid-modifying agents were the United States (40,902 standard units/1000 inhabitants), Greece (39,827 standard units/1000 inhabitants), and France (39,751 standard units/1000 inhabitants). By 2018, Greece, Portugal, and Belgium, were the greatest consumers of lipid-modifying agents (Supplementary Table 3). Over the most recent 5-year period, 64 (95.5%) of the IQVIA countries had increased use of lipid-modifying agents (Supplementary Table 3). During this time period, Belgium, Chile, Ecuador, France, Hungary, Luxembourg, the United States, and Venezuela all experienced decreased consumption of lipid-modifying agents.

When comparing 2008 to 2018, 17 countries (25%) had decreases in statin market share (Figure 4), while statin market share increased in the remainder. Large increases in statin market share occurred in several countries from different regions, such as the Russian Federation, Venezuela, French West Africa, India, China, and the United States. In 2018, subregions with the highest proportion of statin use included Eastern, Northern, and Southern Europe, Western Africa, and Central Asia (Figure 4B).

**Figure 4.**
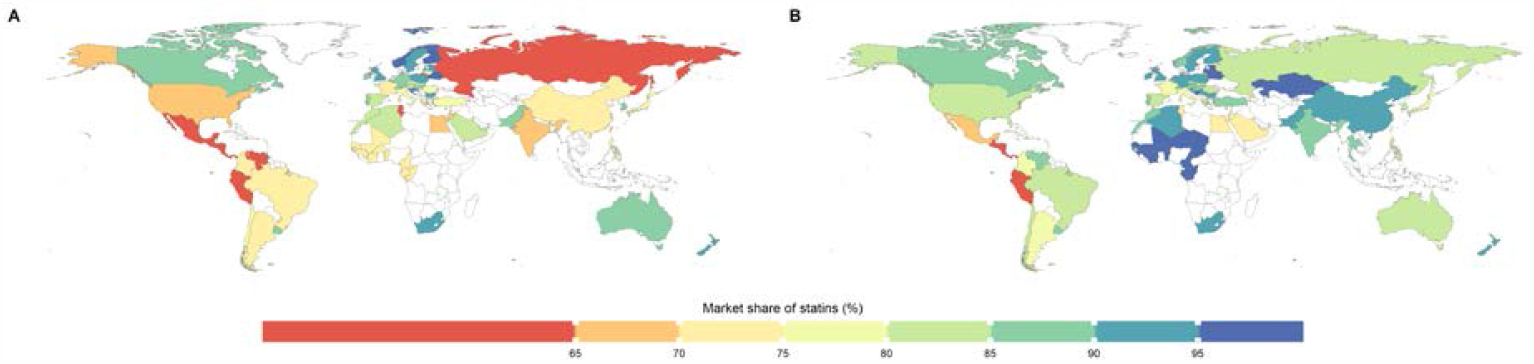
Market Share of Statins by Country in 2008 and 2018. **A**, market share of statins in 2008. **B**, market share of statins in 2018. Market share was calculated as the proportion of statin standard units to all lipid-modifying agent standard units.

### Relationship to Cholesterol Concentrations

In 2018, there was a moderate positive correlation between lipid-modifying agent use and mean total cholesterol concentrations in men (Figure 5A) and women (Figure 5B). However, there was weaker evidence of a correlation for non–HDL cholesterol concentrations (Figure 5C and Figure 5D). Distinct country-level patterns were observed when consumption of lipid-modifying agents and sex-specific total (Figure 6) and non–HDL cholesterol concentrations were compared in 2008 and 2018 (Figure 7). Many countries, such as Korea, Greece, and Puerto Rico, experienced large changes in consumption and small changes in total and non–HDL cholesterol. Others, such as French West Africa, China, and the Philippines, saw little change in the use of lipid-modifying agents, but had increases in cholesterol levels. In contrast, France, and the United States experienced both reductions in use of lipid-modifying agents and total and non–HDL cholesterol concentrations.

**Figure 5.**
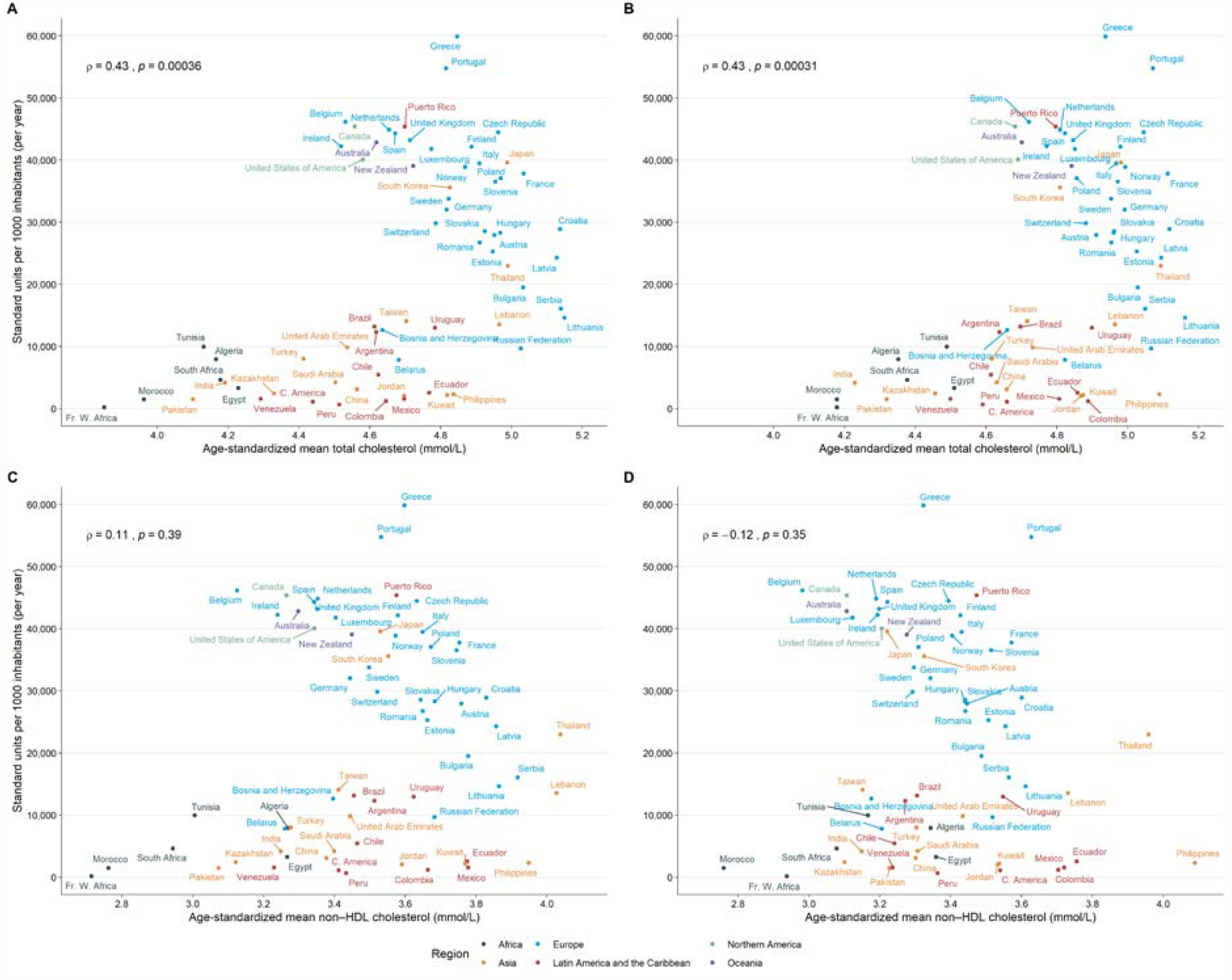
Relationship Between Consumption of Lipid-Modifying Agents and Mean Concentrations of Total Cholesterol and non–HDL Cholesterol in 2018. **A**, consumption of lipid-modifying agents vs age-standardized mean total cholesterol for men. **B**, consumption of lipid-modifying agents vs age-standardized mean total cholesterol for women. **C**, consumption of lipid-modifying agents vs age-standardized non–HDL cholesterol for men. **D**, consumption of lipid-modifying agents vs age-standardized mean non–HDL cholesterol for women. Cholesterol data are from the NCD Risk Factor Collaboration.(6) *ρ* indicates Spearman’s correlation coefficient.

**Figure 6.**
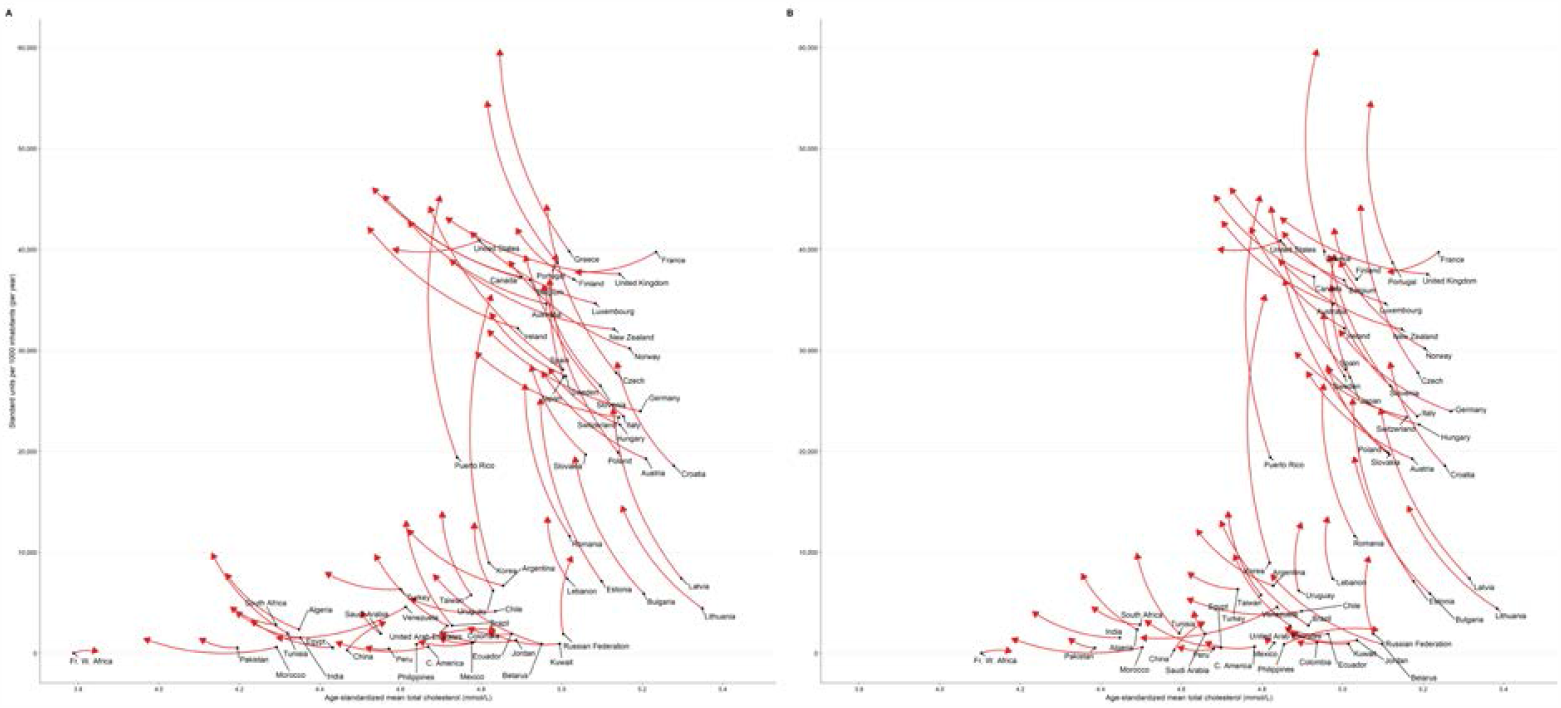
Changes in Lipid-modifying Agent Consumption and Mean Total Cholesterol Concentrations from 2008 to 2018. The start of the arrow (black point) indicates 2008 and the head of the arrow indicates 2018. **A**, consumption of lipid-modifying agents vs age-standardized mean total cholesterol for men. **B**, consumption of lipid-modifying agents vs age-standardized mean total cholesterol for women. Cholesterol data are from NCD Risk Factor Collaboration.(6) Bosnia, Kazakhstan, Netherlands, Serbia, and Thailand do not have MIDAS data for 2008 and were excluded.

**Figure 7.**
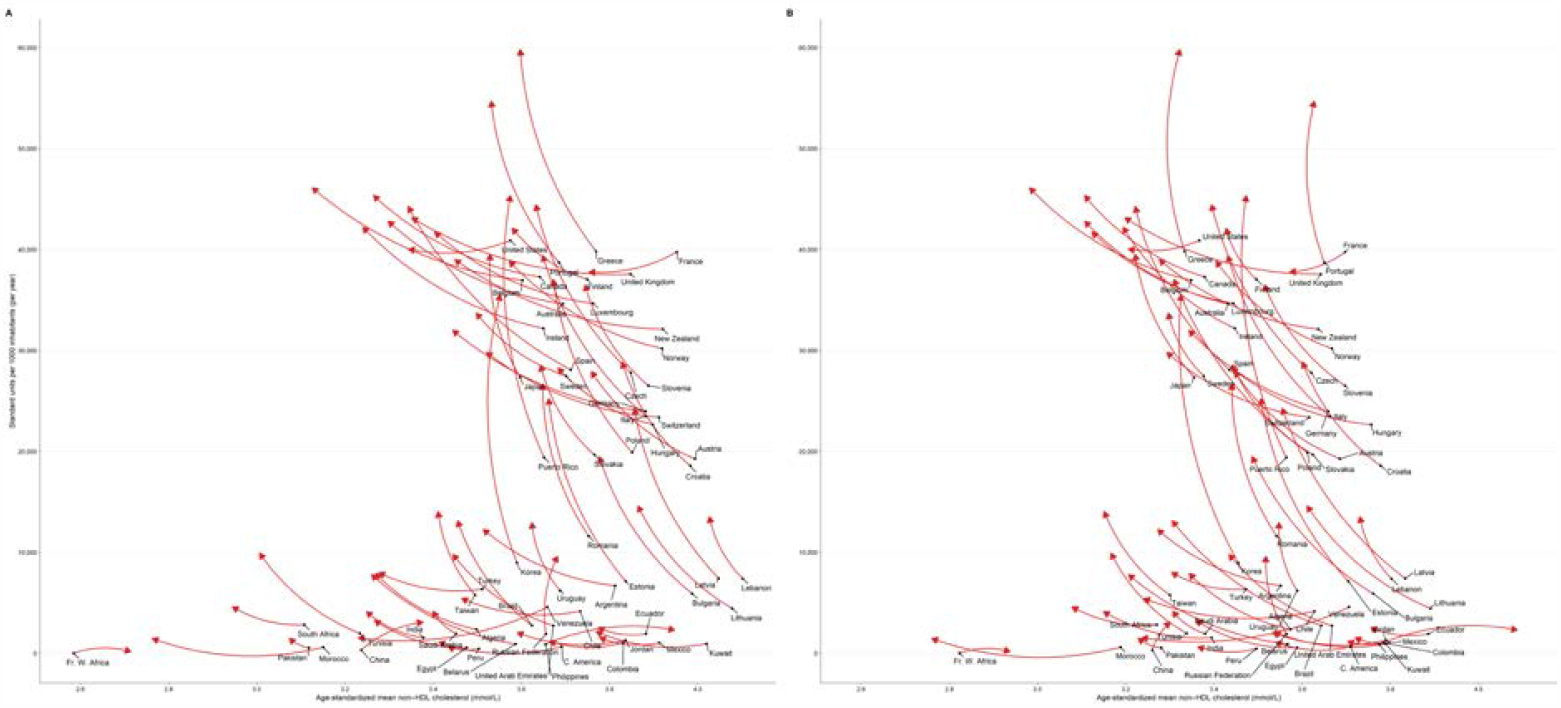
Changes in Lipid-modifying Agent Consumption and Mean non–HDL Cholesterol Concentrations from 2008 to 2018. The start of the arrow (black point) indicates 2008 and the head of the arrow indicates 2018. **A**, consumption of lipid-modifying agents vs age-standardized mean non–HDL cholesterol for men. **B**, consumption of lipid-modifying agents vs age-standardized mean non–HDL cholesterol for women. Cholesterol data are from NCD Risk Factor Collaboration.(6) Bosnia, Kazakhstan, Netherlands, Serbia, and Thailand do not have MIDAS data for 2008 and were excluded.

## DISCUSSION

Our analysis shows a consistent increase in the use of lipid-modifying agents from 2008 to 2018, largely driven by the increased global consumption of statins. In the latter five-year period, rapid uptake of LDL-C lowering nonstatin treatments–PCSK9 inhibitors and ezetimibe–contrasts with declines in consumption of older nonstatin treatments (i.e., fibrates, nicotinic acid, and bile acid sequestrants). Distinct country-level patterns of changes in lipid-modifying agent use and lipid concentrations exist. Although statins have robust evidence of efficacy and safety from multiple randomized controlled trials and are a mainstay of lipid-lowering treatment, there was great diversity in statin market share between countries.

### Comparison with Previous Studies

Few data are available on lipid-modifying agent use, and the market share of statins, in countries outside Northern America and Western Europe. The use of ezetimibe, fibrates, and niacin increased steadily in the United States and Canada from 2002 to 2009.(16–18) In these studies, the overall use of nonstatins was higher in the United States than Canada and is reflected in our findings which saw an increased statin market share in the United States, but a relatively stable market share for statins in Canada. Increases in statin use and the use of generic statins occurred in the United States from 2002 to 2013.(19). Previous reports from European countries describe consistent increases in consumption of lipid-modifying agents.(20–23) Statins were the predominant lipid-modifying agent although the market share of fibrates remained strong in France (∼25%), Belgium (∼25%) and Germany (∼10%) in 2003.(20) Sabo et al reported lower use of statins in Serbia compared with Scandinavian countries.(24) Roth et al reported cross-country comparisons of the proportion of the population treated with lipid-modifying agents and included data from three middle-income countries.(25) While in India, growth in statin prescriptions and a large proportion of combination lipid-modifying agent product use was reported between 2006 to 2010.(26) In East Asia, increased use of statins has been described in Hong Kong and Taiwan.(27,28) Taken together, the global trends in increased use of lipid-modifying agents continued over the past decade, but appear to have weakened since 2013. In contrast to earlier reports from North America, recent trends point to an overall decreased use of fibrates and niacin.

### Changes in Consumption Versus Trends in Cholesterol

The epidemiological transition of elevated blood cholesterol in several developing regions has now been quantified.(6) Rapid growth in the uptake of lipid-modifying agents has coincided with increases in cholesterol concentrations, particularly in countries located in East and Southern Asia, and Northern and Western Africa. However, lipid-modifying agent use in some countries has not kept pace with increases in total and non–HDL cholesterol. Statin use appears to be responsible for about one-third of reductions in total cholesterol in the United States and the United Kingdom.(29,30) For the countries in our analysis, it appears that country-level changes in non–HDL cholesterol over time do not necessarily coincide with a corresponding change in lipid-modifying agent consumption. Several countries had reductions in non–HDL cholesterol and had similar or reduced use of lipid-modifying agents from 2008 to 2018. The patterns observed in this study, support the assertion that declines in non–HDL cholesterol are mostly due to dietary changes, and not increased national use of lipid-modifying agents.(30–33)

### Cholesterol Guidelines and Trials

The timing of publication of key cholesterol guidelines and clinical trials aligns with the observed consumption trends for niacin, ezetimibe, and PCSK9 inhibitors. Evidence of the futility of adding niacin to statin therapy, led to the early stopping of the AIM-HIGH study (published December 2011).(34) In our study, the use of niacin gradually increased and peaked in 2010-2011. Declining use of niacin further accelerated (2013-2018 CAGR −6.92%) after niacin-laropiprant failed to reduce the incidence of major vascular events in the HPS2-THRIVE study (published July 2014).(35) The use of ezetimibe, in contrast, decreased up until 2015, because of the results of two clinical trials which suggested minimal evidence of benefit when ezetimibe was added to a statin: ENHANCE (published April 2008),(36) and SEAS (published September 2008).(37) The resumption of increased use of ezetimibe since 2015 was likely influenced by the evidence of benefit of ezetimibe-simvastatin in the IMPROVE-IT study (published June 2015),(38) the introduction of generic ezetimibe (available in the United States in 2016),(39) and the recommendation to add ezetimibe to statins in high-risk individuals in the 2016 ESC/EAS cholesterol guideline.(40) Interestingly, ezetimibe’s U shaped trajectory means that consumption in 2018 was nearly unchanged from that in 2008. Use of PCSK9 inhibitors increased rapidly after their introduction in 2015, and their use was further supported by evidence of benefit from the FOURIER (published May 2017)(41) and the ODYSSEY OUTCOMES (published November 2018) trials.(42) Consumption of PCSK9 inhibitors will likely continue to increase beyond 2018 given the emphasis on lower LDL cholesterol goals, particularly in very high-risk patients in both the ACC/AHA 2018(2) and EAS/ESC 2019(1) cholesterol guidelines, and potential further price reductions.(43)

### Strengths and Limitations

We have identified diverse country-level trajectories by linking data on national lipid concentrations and lipid-modifying agent consumption. Another strength of our study is that it includes recent data from 83 countries, from different regions and with different income levels. To our knowledge, IQVIA MIDAS is the only platform which permits timely global and cross-country comparisons of medication consumption using locally collected data. Previous studies of lipid-modifying agents have tended to focus only on statin consumption, but we report consumption of all major classes of statin and nonstatin lipid-modifying agents. In particular, a current assessment of the recent trends in use of ezetimibe, PCSK9 inhibitors, and omega-3 fatty acids is needed since emerging evidence and clinical guidelines now suggest their use in select high-risk patients. Our study also has limitations. Comparisons between regions, subregions, and countries should be interpreted in the context of the available data and total market coverage of the included countries. Less than 100% market coverage underestimates the true consumption of lipid-modifying agents (e.g. Korea lacks hospital data). However, total pharmaceutical market coverage in most countries was greater than 80%. Lipid-modifying agents are taken on a chronic basis and are generally obtained from retail stores in countries that are not centered on the delivery of healthcare in hospitals; retail consumption data was available for all countries except for Taiwan and Thailand. Conclusions from our ecological study should not be interpreted as being applicable to individual inhabitants of each country. Furthermore, relationships between changes in lipid-modifying agent consumption and total and non–HDL cholesterol are not causal. We did not adjust for other factors that may influence lipid-modifying agent consumption such as indication, access to healthcare services, affordability of medications, and dietary patterns. However, distinct country-level patterns are useful for generating hypotheses for further research.

## Conclusion

Growth in the use of lipid-modifying agents has continued globally and statins are the dominant class of lipid-modifying agent. However, three decades since the introduction of the first statin, their adoption in Africa, Asia, and Latin America and the Caribbean remain markedly lower than in Northern America and Europe, but is growing rapidly.

## Supporting information

Strobe checklist

## Data Availability

The underlying MIDAS data were provided by IQVIA under license. The terms of our agreement do not permit disclosure, sublicensing, or sharing of IQVIA MIDAS data.
The underlying cholesterol data are available from the NCD Risk Factor Collaboration.

http://www.ncdrisc.org/data-downloads-cholesterol.html

## Author Contributions

Conceptualization: Blais, Yap, Chan

Formal Analysis: Blais, Y. Wei

Funding acquisition: Chan, Wong

Resources: Chan, Wong, L. Wei

Visualization: Yap, Blais

Supervision: Chan, Wong

Writing - original draft: Blais, Yap

Writing - review and editing: All authors

## Data Availability

The underlying MIDAS data were provided by IQVIA under license. The terms of our agreement do not permit disclosure, sublicensing, or sharing of IQVIA MIDAS data.

The underlying cholesterol data are available from the NCD Risk Factor Collaboration: http://www.ncdrisc.org/data-downloads-cholesterol.html.

## SUPPLEMENTARY MATERIAL

**Supplementary Table 1.**
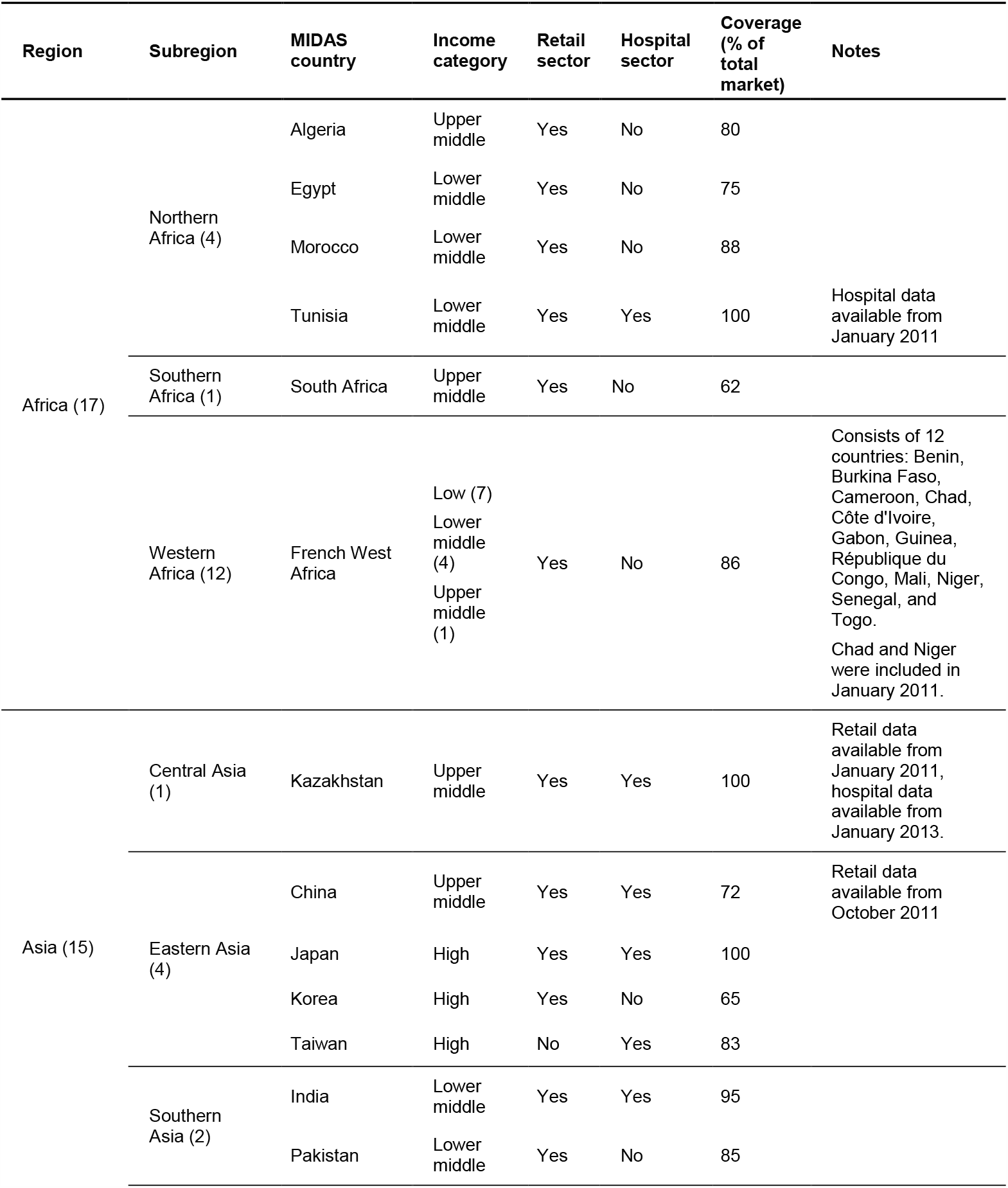
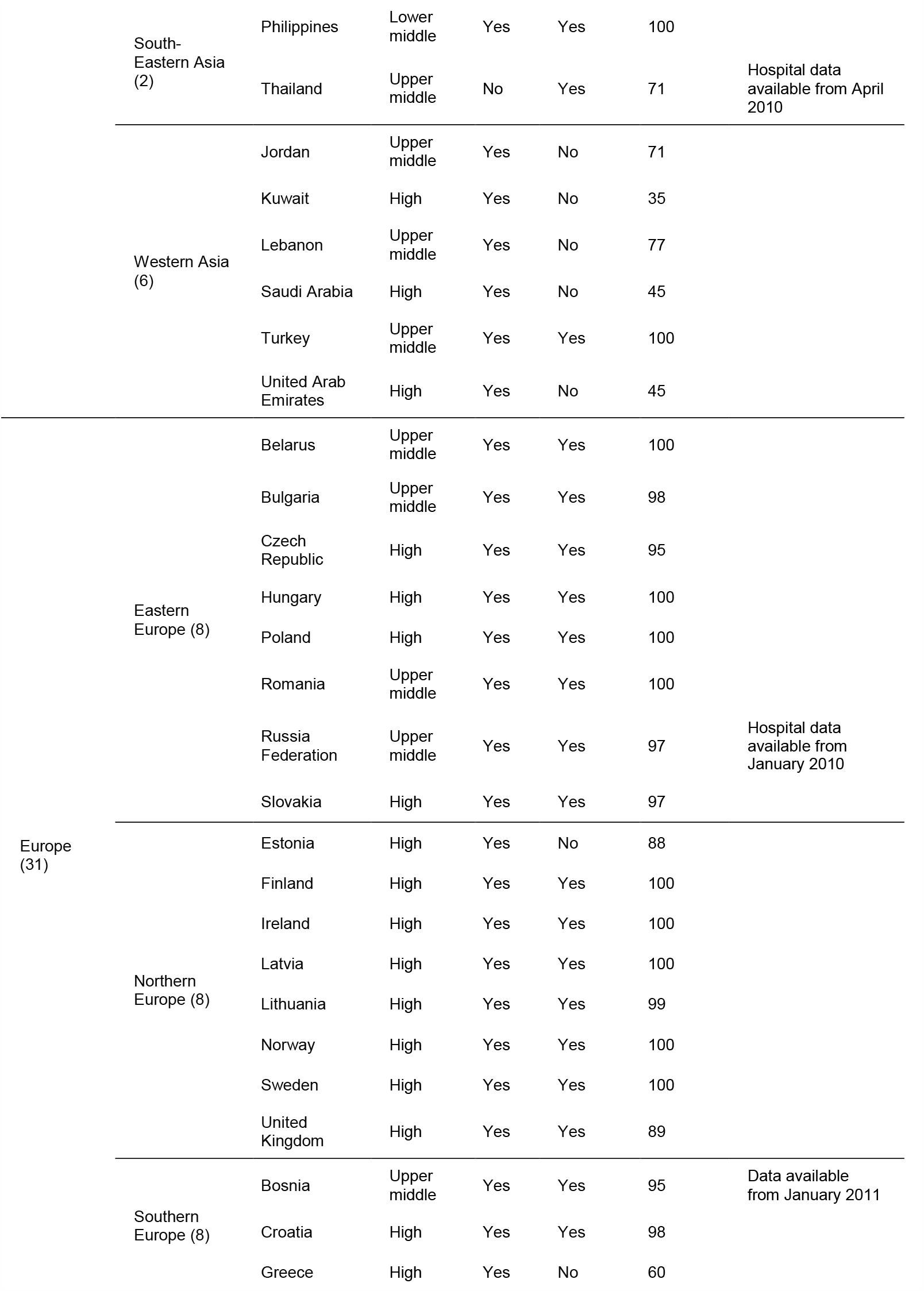
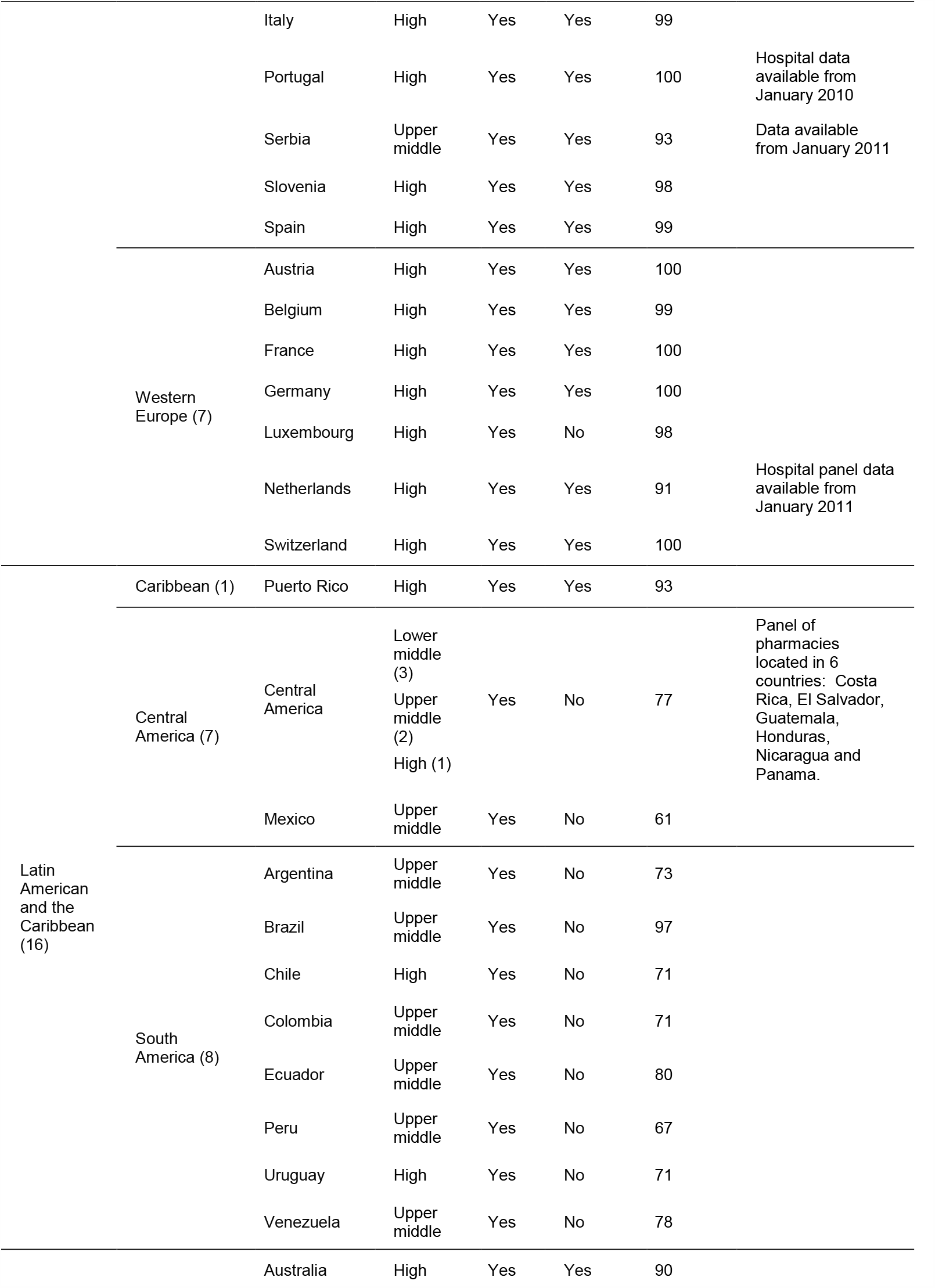
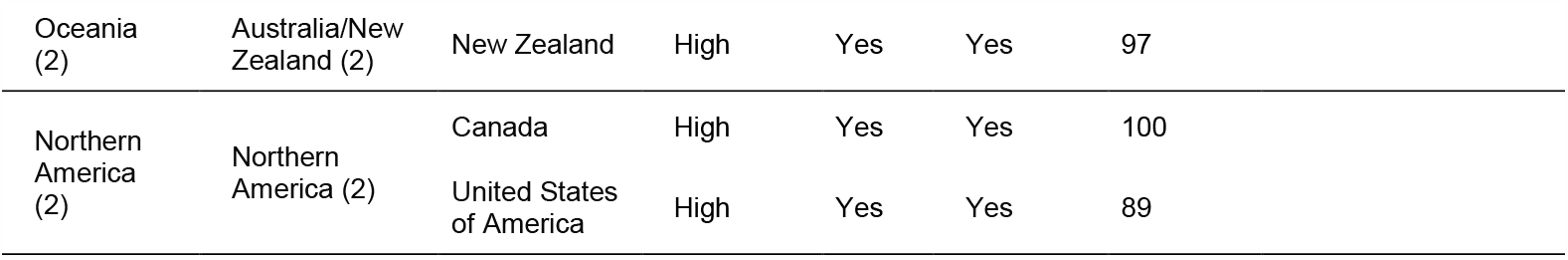
Geographic Classification, Income Category, and Total Pharmaceutical Market Data Coverage of the included IQVIA MIDAS Countries. Channels (retail and hospital) in each country that do not have 100% audit coverage are projected to 100% of each channel by IQVIA. Coverage of MIDAS country panels indicates data were available for analysis in this study. Figures in brackets indicate the number of countries included in each region, subregion, or income category. Income category was defined according to the World Bank Fiscal Year 2020 (calendar year 2018).

**Supplementary Table 2.**
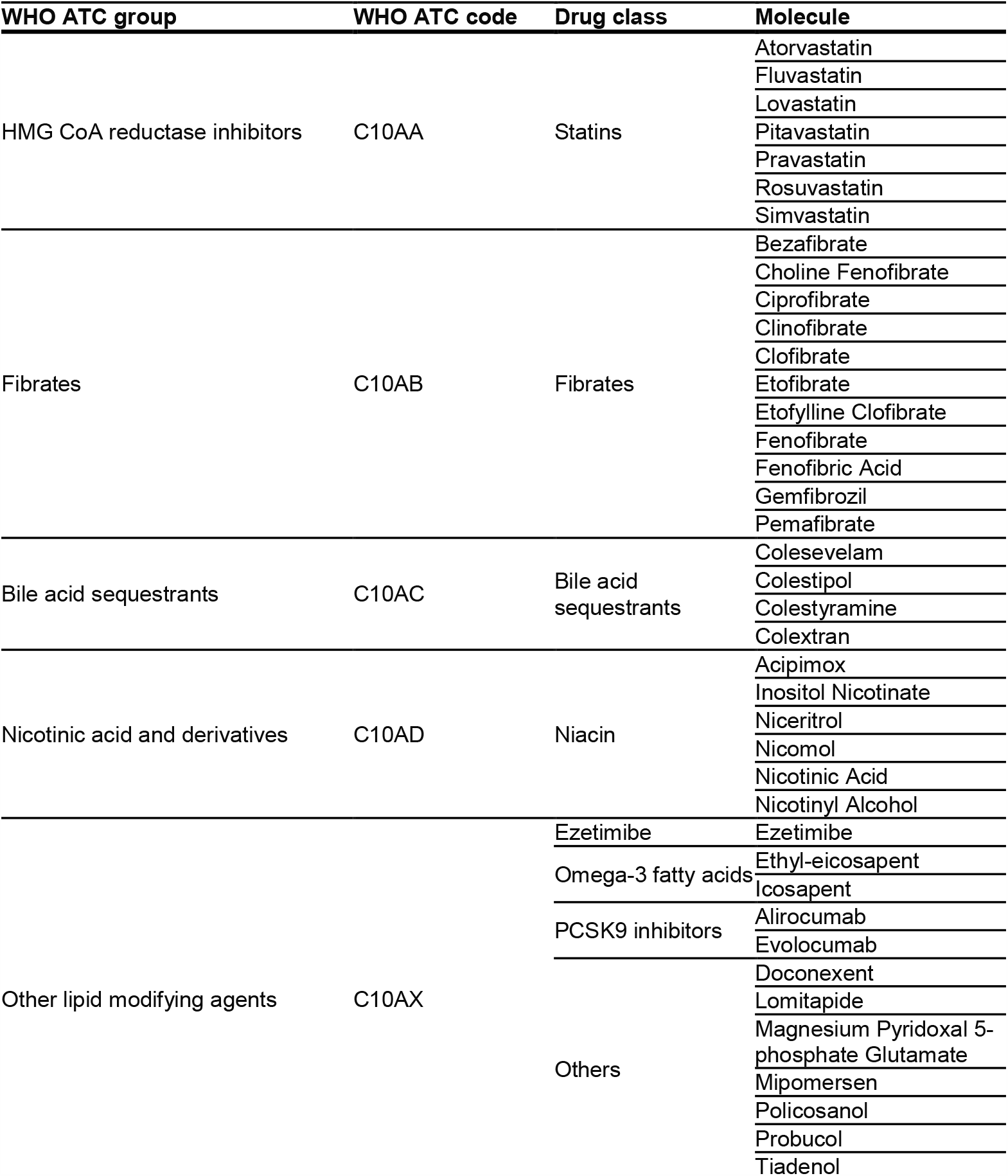
WHO Anatomic Therapeutic Chemical Groups, Codes, Agent Classes, and Medicines Included in this Study.

**Supplementary Table 3.**
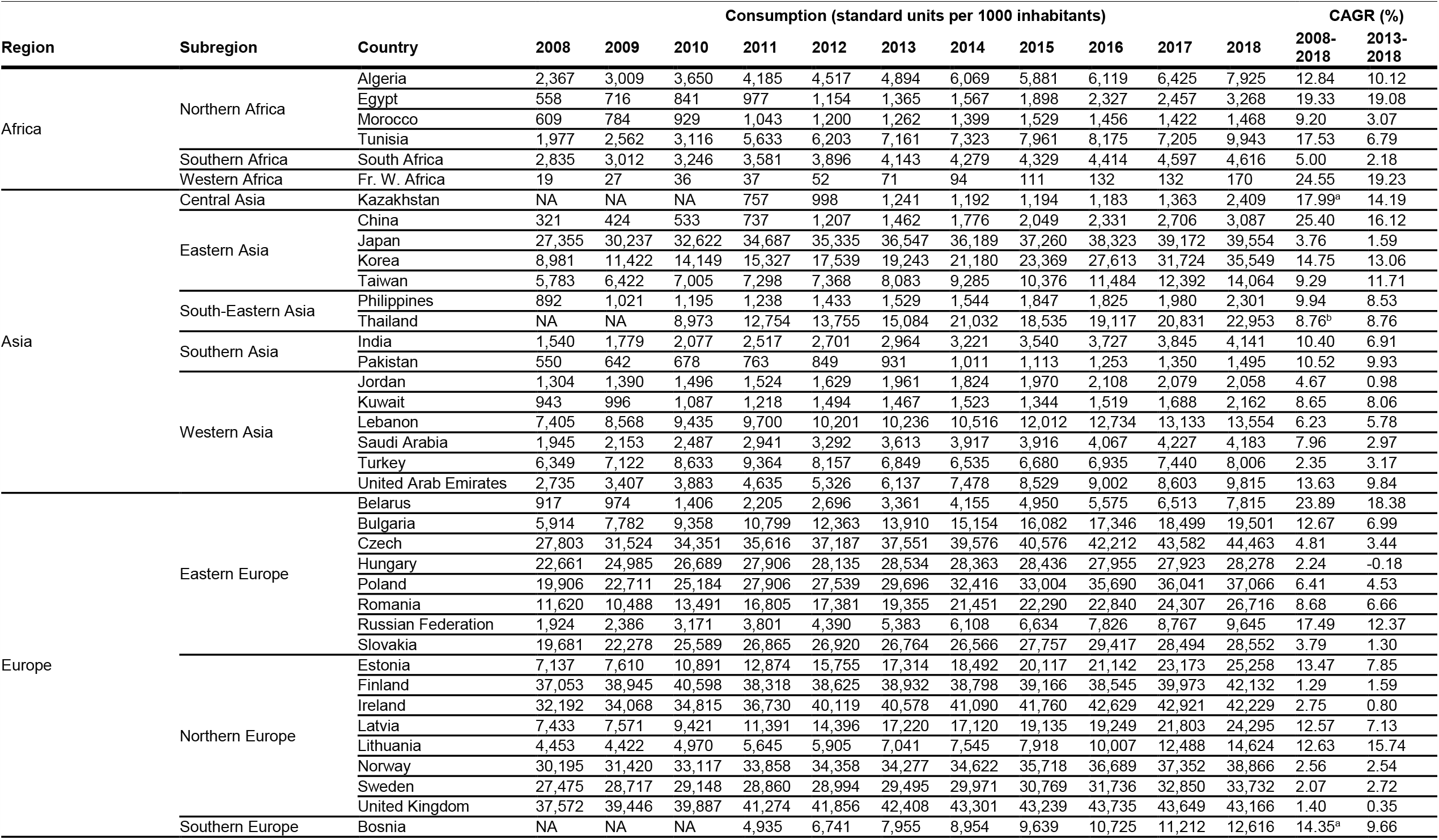
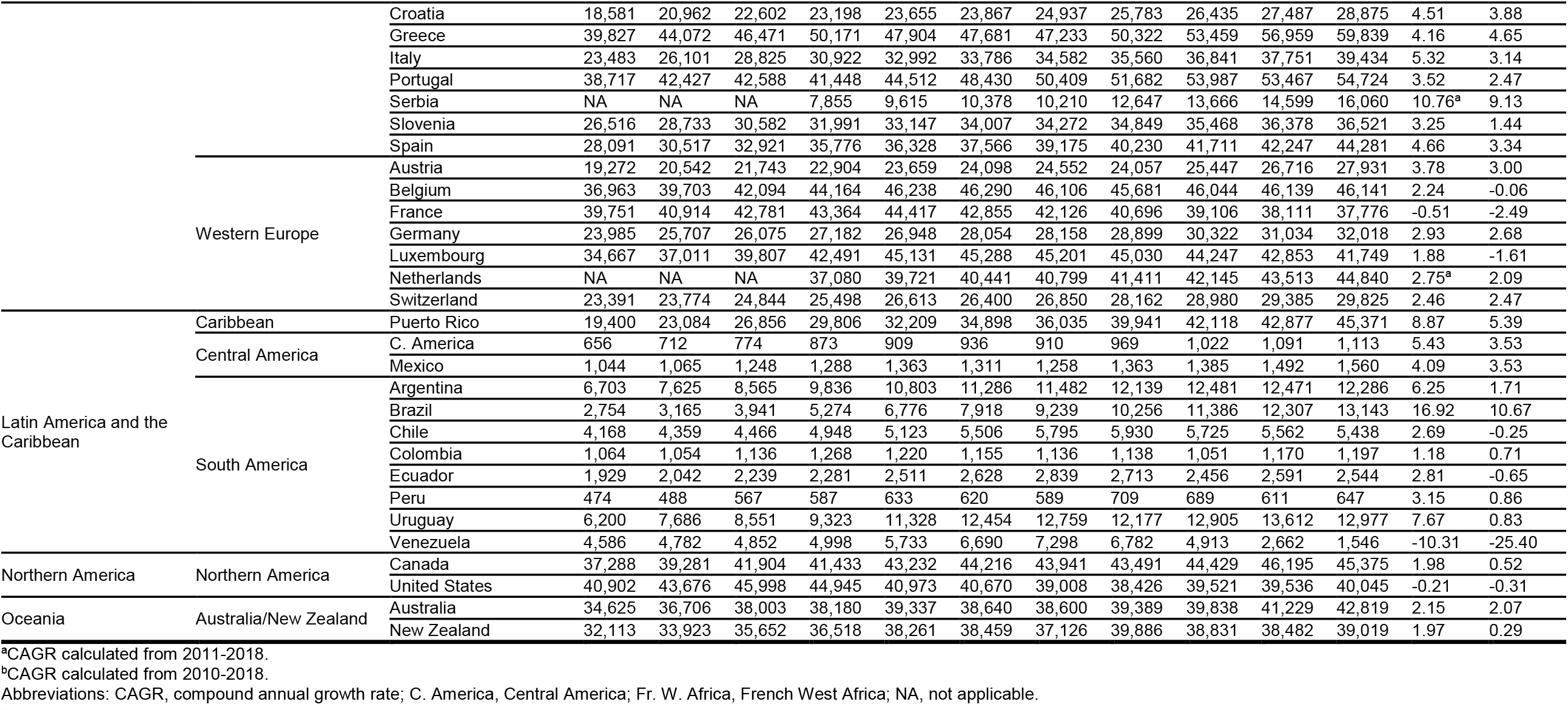
Annual Use of All Lipid-Modifying Agents in Each MIDAS Country from 2008 to 2018.

